# Orchestrated multi agents sustain accuracy under clinical-scale workloads compared to a single agent

**DOI:** 10.1101/2025.08.22.25334049

**Authors:** Eyal Klang, Mahmud Omar, Ganesh Raut, Reem Agbareia, Prem Timsina, Robert Freeman, Nicholas Gavin, Lisa Stump, Alexander W Charney, Benjamin S Glicksberg, Girish N Nadkarni

## Abstract

We tested state-of-the-art large language models (LLMs) in two configurations for clinical-scale workloads: a single agent handling heterogeneous tasks versus an orchestrated multi-agent system assigning each task to a dedicated worker. Across retrieval, extraction, and dosing calculations, we varied batch sizes from 5 to 80 to simulate clinical traffic. Multi-agent runs maintained high accuracy under load (pooled accuracy 90.6% at 5 tasks, 65.3% at 80) while single-agent accuracy fell sharply (73.1% to 16.6%), with significant differences beyond 10 tasks (FDR-adjusted p < 0.01). Multi-agent execution reduced token usage up to 65-fold and limited latency growth compared with single-agent runs. The design’s isolation of tasks prevented context interference and preserved performance across four diverse LLM checkpoints. This is the first evaluation of LLM agent architectures under sustained, mixed-task clinical workloads, showing that lightweight orchestration can deliver accuracy, efficiency, and auditability at operational scale.

## Main

Large-language models (LLMs) are already used in clinical research and practice because they can read free text, reason over symptoms and generate fluent advice ^1^. Recent work wraps these models in autonomous “agents” that decide, for example, which database query or dose calculator to invoke at each step, reflecting the multi-step structure of clinical problem-solving ^2^. AMIE, published in *Nature*, showed that a single dialogue-optimised LLM can match physicians in simulated consultations ^3^. That study, however, evaluated one case at a time and did not examine how accuracy or cost change when workloads grow or when tasks require many different tools at the same time.

We conduct the first study to test state-of-the-art LLM agents in two execution modes. In the single-agent baseline one agent receives an entire batch of heterogeneous tasks. In the orchestrated mode with multiple agents a lightweight orchestrator sends each task to a dedicated worker agent, which calls one domain tool before returning its answer for aggregation. Batch sizes ranged from□5□to□80 tasks to mimic clinical traffic. Tasks fell into three categories: Retrieval (find PubMed abstracts), Extraction (pull fields from a discharge note), and Dosing (solve a medication⍰math calculation) (tasks are detailed in **Supplement Section 1**). This let us test whether task⍰level delegation preserves accuracy and limits latency and token cost as conversational agents scale, while keeping every reasoning step transparent and traceable (**Figure 1**).

**Figure 1.**
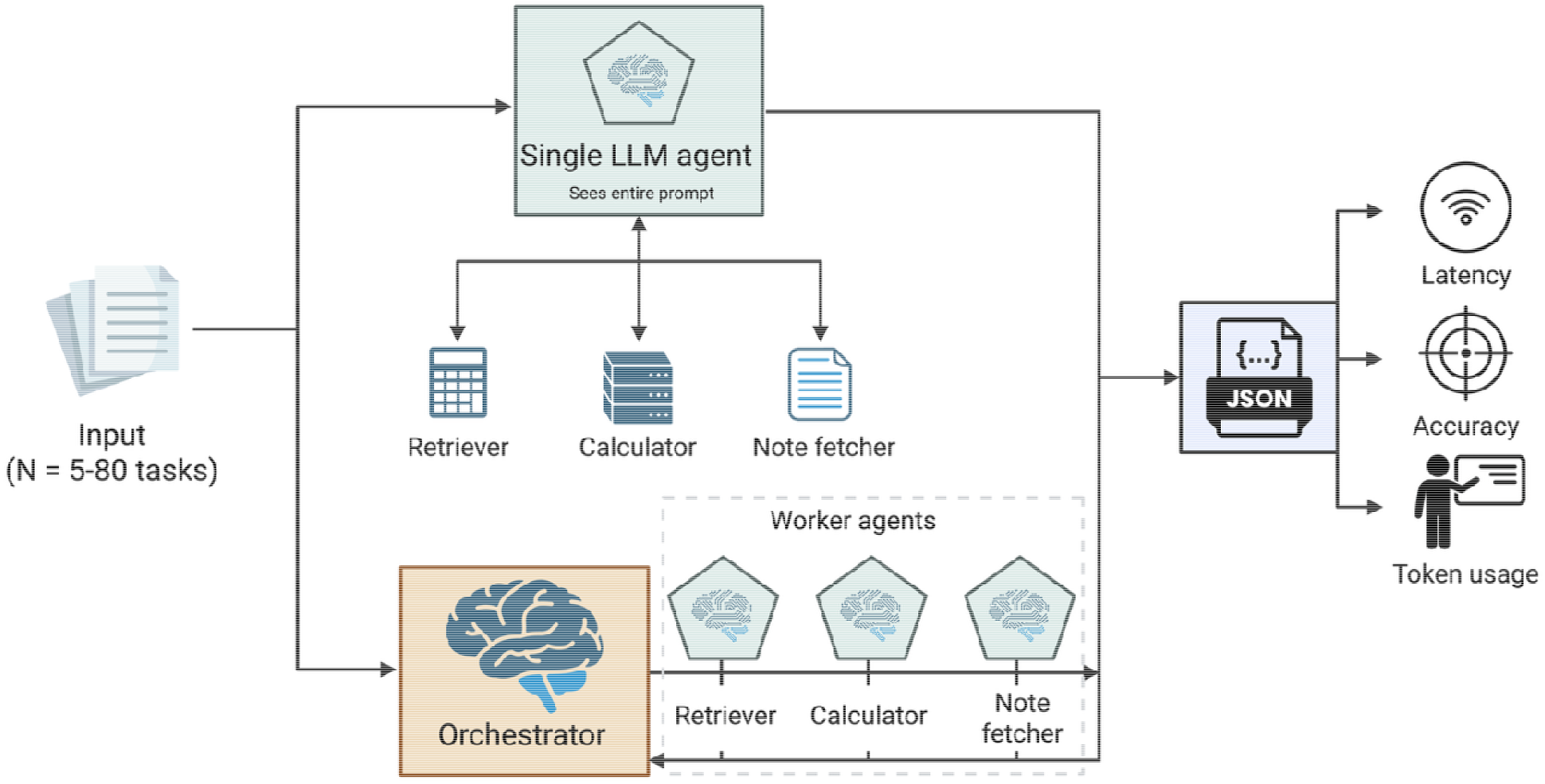
Overview of the pipeline design.

In our tests, accuracy diverged once batch size passed ten tasks (Figure□2). When results were pooled, multi⍰agent runs held 90.6□% at batches of five tasks and 65.3□% at eighty (95□%□CI□56–74□%), whereas single⍰agent accuracy fell from 73.1□% to 16.6□%. GPT⍰4.1⍰mini illustrates the gap: its multi⍰agent accuracy stayed between 96□% and 91.4□% across all batch sizes, while the single agent declined from 96□% to 33.9□%. GPT⍰4.1⍰nano, Llama⍰2⍰70B and Qwen⍰3⍰8B showed the same pattern, with single⍰agent accuracy dropping far below their multi⍰agent counterparts. All differences between topologies beyond ten tasks were significant (Welch□t⍰test, FDR⍰adjusted p□<□0.01). Detailed values are listed in **Supplement**□**Section**□**1**.

**Figure 2.**
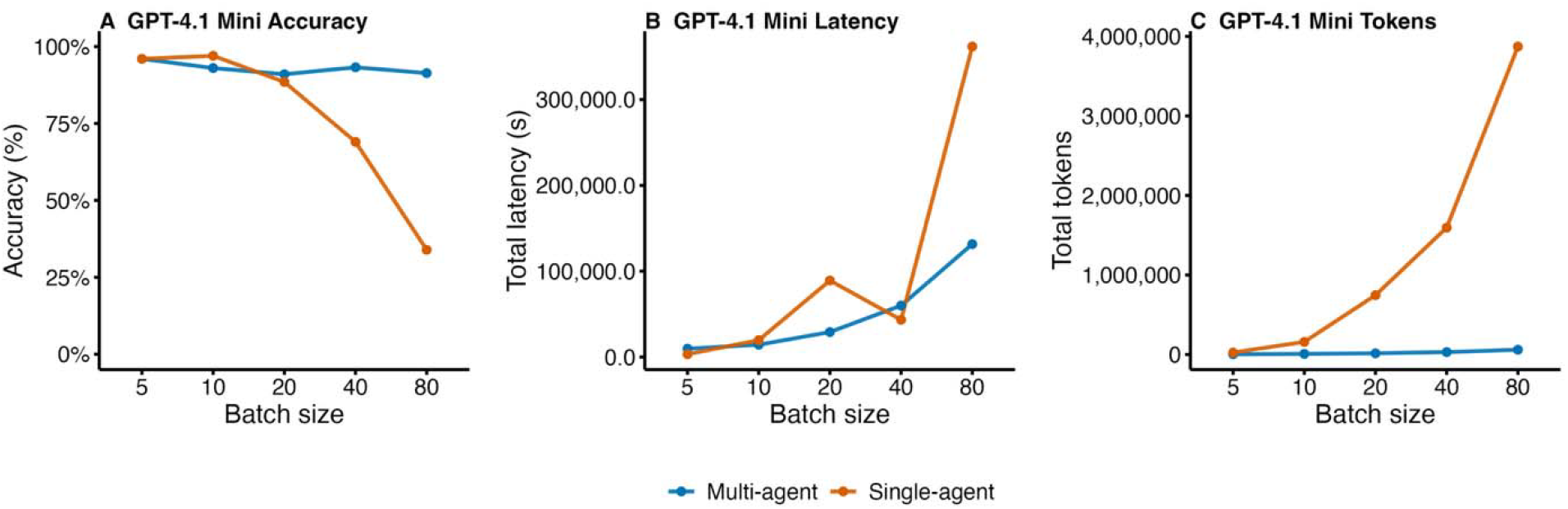
Accuracy and token usage across: above-GPT-4.1 mini model, while below represents the pooled data across the 4 models.

Token growth differed sharply between agent types.□For GPT⍰4.1⍰mini the multi⍰agent batch rose only from□3.7□k tokens at five tasks to□60□k at eighty, whereas the single agent expanded from□25□k to□3.9□M—a 65⍰fold gap at the highest load.□GPT⍰4.1⍰nano showed the same direction but at higher totals.□Llama⍰2⍰70B and Qwen⍰3⍰8B were less consistent; many large single⍰agent batches for these models failed early and returned no JSON, confounding direct comparison.

Total latency rose with batch size in both modes, but the single⍰agent curve climbed faster. For GPT⍰4.1⍰mini, multi⍰agent batches grew from□9□728□s at five tasks to□131□603□s at eighty, whereas single⍰agent batches rose from□3□258□s to□361□864□s. The other models showed the same direction: multi⍰agent latency increased predictably, while single⍰agent runs jumped higher and displayed wider confidence intervals; several large single⍰agent batches for Qwen⍰3⍰8B and Llama⍰2⍰70B failed and returned no JSON.

Delegating each task to its own worker appears to insulate the LLM from context interference, so accuracy remains high even when many unrelated prompts arrive at once. This insulation likely stems from two linked mechanisms. First, each worker receives only the tokens that matter for a single decision, so attention is not diluted across irrelevant material ^4^. Second, the orchestrator re-assembles answers without expanding the prompt seen by any one model call, keeping the effective context well within the range the model was tuned to handle ^5^. These benefits come almost for free: the orchestrator adds minimal latency and no gradient updates, yet it may spare many millions of tokens when workloads rise to clinical or enterprise scale.

Model scale in this pipeline appears to drive stability. The larger the checkpoint, the longer accuracy persists under multi-agent load. GPT-mini, likely the most parameter-rich model in the ensemble, is by far the best performer and barely drifts from its initial high accuracy. GPT-nano, consistent with its smaller size, begins to erode once batch size rises. Llama-2-70B follows the same pattern and sheds more accuracy as batches grow. As expected from this trend, Qwen-3-8B, with the smallest parameter budget and limited exposure to mixed-tool prompts, degrades fastest.

This is the first study to show that a lightweight orchestrator can turn existing LLMs into reliable, token-efficient teams for complex clinical workflows. By dividing labour into single-tool subtasks, the architecture keeps accuracy high even when many requests arrive simultaneously-an essential property for automated, large-scale use. Recent agent studies in medicine, AMIE for diagnostic dialogue ^3^, DRAGON for radiology reporting ^6^, and a handful of retrieval-augmented chart summarizers, each examined one model, one patient at a time, and did not examine performance, cost or latency under clinical scale loads ^7^. Our experiments address a different but linked and practical aspect: context overload when dozens of heterogeneous tasks arrive together. The findings show that an orchestrator-worker layout sustains accuracy, tokens, and latency in a regime where single-prompt agents collapse. This benefit holds across four checkpoints with widely different sizes and pre-training recipes, suggesting the gain comes from task partitioning rather than from any model-specific optimisation. By constraining each worker to a single tool and a short prompt, the design also produces a transparent audit trail, as every intermediate call can be logged and replayed, addressing a key regulatory concern that earlier end-to-end agents leave unresolved.

Our multi-agent approach therefore offers a practical, and transformative path to clinical decision support that is both affordable and auditable. Future studies should test similar pipelines within specific specialties, and against real patient records to establish generalisability and bedside utility.

## Methods

### Experimental protocol and agentic pipelines

We tested two agent layouts on three task classes: retrieving oncology abstracts, extracting fields from discharge notes and solving numeric dosing problems. The retrieval set is a FAISS index of 234,650 PubMed records on neoplasms (2020-01-01 to 2025-05-30) embedded with GIST-large. The extraction set contains 331,793 electronic health record (EHR) summaries with admission and discharge dates jittered across 2005-2019 ^8^. The calculation set is generated by 20 templates that cover routine weight-, surface- and clearance-based dosing. Ground truth is a single canonical string per item; accuracy is the proportion of exact matches after case- and whitespace normalisation (More detailed description is provided in the **Supplement Section 2**).

Agents could call a k-NN retriever, a sandboxed calculator and a note fetcher. In the single-agent condition one language model received the full batch of N tasks and up to 10 × N turns. In the multi-agent condition an orchestrator dispatched each task to one worker, each worker handled one tool, and the orchestrator merged the replies. Four instruction-tuned checkpoints ran under both topologies with random seed 42.

Batch sizes were 5, 10, 20, 40 and 80; ten non-overlapping batches were generated for every model-topology pair. Open-weight models used an 8 × H100-80 GB node; hosted models were called through vendor APIs. We recorded wall-clock latency from the first call to final JSON and counted total input + output tokens.

## Data Availability

All data produced in the present study are available upon reasonable request to the authors

## Statistical analysis

Accuracy was evaluated with two-sided Welch t-tests; latency and token totals with Wilcoxon rank-sum tests. P values were Benjamini–Hochberg corrected within each metric family. Analyses ran in R 4.3 and full code and logs are supplied in the Supplement.

## Abbreviations

LLM: Large Language Model
PED: ediatric Emergency Department
GT: Ground Truth
CI: Confidence Interval
pp: Percentage Points.

## Notes

**Financial disclosure** – This work was supported in part through the computational and data resources and staff expertise provided by Scientific Computing and Data at the Icahn School of Medicine at Mount Sinai and supported by the Clinical and Translational Science Awards (CTSA) grant UL1TR004419 from the National Center for Advancing Translational Sciences. Research reported in this publication was also supported by the Office of Research Infrastructure of the National Institutes of Health under award number S10OD026880 and S10OD030463. The content is solely the responsibility of the authors and does not necessarily represent the official views of the National Institutes of Health. The funders played no role in study design, data collection, analysis and interpretation of data, or the writing of this manuscript.

### Competing Interest Statement

The authors have declared no competing interest.

### Funding Statement

Financial disclosure: This work was supported in part through the computational and data resources and staff expertise provided by Scientific Computing and Data at the Icahn School of Medicine at Mount Sinai and supported by the Clinical and Translational Science Awards (CTSA) grant UL1TR004419 from the National Center for Advancing Translational Sciences. Research reported in this publication was also supported by the Office of Research Infrastructure of the National Institutes of Health under award number S10OD026880 and S10OD030463. The content is solely the responsibility of the authors and does not necessarily represent the official views of the National Institutes of Health. The funders played no role in study design, data collection, analysis and interpretation of data, or the writing of this manuscript.

