## Supplement for "Orchestrated multi agents sustain accuracy under clinical-scale workloads compared to a single agent"

**Section 1: full results.**

Column definitions for the following tables:

| Column | Meaning |
|---|---|
| dataset | Model checkpoint evaluated (for example, *gpt_mini* or *llama-2-70B*). |
| type | Agent topology: *multi* = multi-agent with orchestrator; *single* = single-agent baseline. |
| N_tasks | Batch size, that is, number of tasks processed together. |
| n | Number of independent batches run for that condition (replicates). |
| mean_acc | Mean accuracy across the *n* batches (exact-match rate). |
| se_acc | Standard error of the mean accuracy. |
| ci_low, ci_high | Lower and upper bounds of the two-sided 95 % confidence interval for the mean accuracy. |

**Table S1. Accuracy by dataset × agent × batch size.**

| dataset | type | N_tasks | n | mean_acc | se_acc | ci_low | ci_high |
|---|---|---|---|---|---|---|---|
| **gpt_mini** | multi | 5 | 10 | 96.0% | 2.7% | 90.0% | 100.0% |
| **gpt_mini** | multi | 10 | 10 | 93.0% | 2.1% | 88.2% | 97.8% |
| **gpt_mini** | multi | 20 | 10 | 91.0% | 2.8% | 84.7% | 97.3% |
| **gpt_mini** | multi | 40 | 10 | 93.3% | 0.7% | 91.6% | 94.9% |
| **gpt_mini** | multi | 80 | 10 | 91.4% | 1.7% | 87.5% | 95.3% |
| **gpt_mini** | single | 5 | 10 | 96.0% | 2.7% | 90.0% | 102.0% |
| **gpt_mini** | single | 10 | 10 | 97.0% | 1.5% | 93.5% | 100.5% |
| **gpt_mini** | single | 20 | 10 | 88.5% | 2.6% | 82.6% | 94.4% |
| **gpt_mini** | single | 40 | 10 | 69.0% | 6.7% | 53.9% | 84.1% |
| **gpt_mini** | single | 80 | 10 | 33.9% | 2.8% | 27.6% | 40.2% |
| **gpt_nano** | multi | 5 | 10 | 82.0% | 6.3% | 67.8% | 96.2% |
| **gpt_nano** | multi | 10 | 10 | 73.3% | 7.9% | 55.4% | 91.2% |
| **gpt_nano** | multi | 20 | 10 | 71.9% | 4.3% | 62.1% | 81.6% |
| **gpt_nano** | multi | 40 | 10 | 71.8% | 3.8% | 63.1% | 80.4% |
| **gpt_nano** | multi | 80 | 10 | 59.3% | 5.2% | 47.4% | 71.1% |
| **gpt_nano** | single | 5 | 10 | 60.0% | 4.5% | 49.9% | 70.1% |
| **gpt_nano** | single | 10 | 10 | 42.2% | 6.1% | 28.5% | 56.0% |
| **gpt_nano** | single | 20 | 10 | 33.8% | 4.5% | 23.5% | 44.0% |
| **gpt_nano** | single | 40 | 10 | 16.3% | 2.0% | 11.6% | 20.9% |
| **gpt_nano** | single | 80 | 10 | 10.4% | 2.7% | 4.1% | 16.6% |
| **llama_xlam_2_70b** | multi | 5 | 10 | 92.0% | 3.3% | 84.6% | 99.4% |
| **llama_xlam_2_70b** | multi | 10 | 10 | 93.0% | 1.5% | 89.5% | 96.5% |
| **llama_xlam_2_70b** | multi | 20 | 10 | 78.9% | 6.5% | 64.3% | 93.5% |
| **llama_xlam_2_70b** | multi | 40 | 10 | 76.9% | 6.2% | 63.0% | 90.9% |

| | | | | | | | |
|---|---|---|---|---|---|---|---|
| **llama_xlam_2_70b** | multi | 80 | 10 | 83.6% | 3.3% | 76.1% | 91.0% |
| **llama_xlam_2_70b** | single | 5 | 10 | 90.0% | 3.3% | 82.5% | 97.5% |
| **llama_xlam_2_70b** | single | 10 | 10 | 91.1% | 1.9% | 86.8% | 95.4% |
| **llama_xlam_2_70b** | single | 20 | 10 | 47.5% | 6.4% | 33.1% | 61.9% |
| **llama_xlam_2_70b** | single | 40 | 10 | 32.1% | 6.8% | 16.7% | 47.4% |
| **qwen_3_8B** | multi | 5 | 10 | 100.0% | 0.0% | 100.0% | 100.0% |
| **qwen_3_8B** | multi | 10 | 10 | 91.0% | 1.8% | 86.9% | 95.1% |
| **qwen_3_8B** | multi | 20 | 10 | 81.5% | 2.9% | 75.0% | 88.0% |
| **qwen_3_8B** | multi | 40 | 10 | 69.7% | 7.2% | 53.5% | 85.9% |
| **qwen_3_8B** | multi | 80 | 10 | 29.0% | 8.1% | 10.6% | 47.4% |
| **qwen_3_8B** | single | 5 | 10 | 25.7% | 4.8% | 14.9% | 36.5% |
| **qwen_3_8B** | single | 10 | 10 | 6.7% | 1.8% | 2.5% | 10.8% |
| **qwen_3_8B** | single | 20 | 10 | 3.0% | 0.9% | 1.0% | 5.0% |
| **qwen_3_8B** | single | 40 | 10 | 1.9% | 0.4% | 1.0% | 2.8% |
| **qwen_3_8B** | single | 80 | 10 | 1.3% | 0.3% | 0.6% | 1.9% |

**Tables S2. Total latency (s) by dataset × agent × batch size.**

| dataset | type | N_tasks | n | mean_val | se_val | ci_low |
|---|---|---|---|---|---|---|
| **gpt_mini** | multi | 5 | 10 | 9,728.00 | 2,335.08 | 4,445.69 |
| **gpt_mini** | multi | 10 | 10 | 14,333.50 | 110.49 | 14,083.56 |
| **gpt_mini** | multi | 20 | 10 | 29,098.80 | 389.74 | 28,217.14 |
| **gpt_mini** | multi | 40 | 10 | 60,002.90 | 327.57 | 59,261.88 |
| **gpt_mini** | multi | 80 | 10 | 131,603.00 | 2,344.26 | 126,299.91 |
| **gpt_mini** | single | 5 | 10 | 3,258.40 | 448.82 | 2,243.10 |
| **gpt_mini** | single | 10 | 10 | 19,641.50 | 6,352.34 | 5,271.51 |
| **gpt_mini** | single | 20 | 10 | 89,159.50 | 22,890.47 | 37,377.66 |
| **gpt_mini** | single | 40 | 10 | 43,344.50 | 12,411.57 | 15,267.57 |
| **gpt_mini** | single | 80 | 10 | 361,863.70 | 179,003.44 | -43,070.21 |
| **gpt_nano** | multi | 5 | 10 | 6,414.50 | 58.66 | 6,281.80 |
| **gpt_nano** | multi | 10 | 10 | 13,251.70 | 513.14 | 12,090.90 |
| **gpt_nano** | multi | 20 | 10 | 28,573.70 | 1,955.27 | 24,150.58 |
| **gpt_nano** | multi | 40 | 10 | 51,440.80 | 1,511.29 | 48,022.03 |
| **gpt_nano** | multi | 80 | 10 | 100,550.70 | 5,647.78 | 87,774.53 |
| **gpt_nano** | single | 5 | 10 | 5,384.70 | 609.12 | 4,006.78 |
| **gpt_nano** | single | 10 | 10 | 7,845.30 | 659.17 | 6,354.16 |
| **gpt_nano** | single | 20 | 10 | 11,743.00 | 1,781.59 | 7,712.76 |
| **gpt_nano** | single | 40 | 10 | 25,231.30 | 8,822.03 | 5,274.48 |
| **gpt_nano** | single | 80 | 10 | 14,181.50 | 4,619.84 | 3,730.70 |
| **llama_xlam_2_70b** | multi | 5 | 10 | 85,567.30 | 3,708.85 | 77,177.29 |
| **llama_xlam_2_70b** | multi | 10 | 10 | 194,376.00 | 3,785.55 | 185,812.49 |
| **llama_xlam_2_70b** | multi | 20 | 10 | 454,734.90 | 39,026.26 | 366,451.36 |

| | | | | | | |
|---|---|---|---|---|---|---|
| **llama_xlam_2_70b** | multi | 40 | 10 | 838,632.60 | 98,746.13 | 615,253.33 |
| **llama_xlam_2_70b** | multi | 80 | 10 | 3,315,372.10 | 1,510,773.36 | -102,234.68 |
| **llama_xlam_2_70b** | single | 5 | 10 | 57,707.30 | 4,482.54 | 47,567.10 |
| **llama_xlam_2_70b** | single | 10 | 10 | 179,635.30 | 11,180.75 | 154,342.69 |
| **llama_xlam_2_70b** | single | 20 | 10 | 312,827.90 | 76,878.36 | 138,916.97 |
| **llama_xlam_2_70b** | single | 40 | 10 | 578,546.30 | 113,230.47 | 322,401.18 |
| **qwen_3_8B** | multi | 5 | 10 | 321,068.90 | 47,051.26 | 214,631.55 |
| **qwen_3_8B** | multi | 10 | 10 | 272,956.10 | 123,639.05 | -6,734.85 |
| **qwen_3_8B** | multi | 20 | 10 | 369,713.40 | 126,273.13 | 84,063.75 |
| **qwen_3_8B** | multi | 40 | 10 | 1,046,207.90 | 621,769.29 | -360,331.95 |
| **qwen_3_8B** | multi | 80 | 10 | 2,036,080.70 | 847,342.92 | 119,257.85 |
| **qwen_3_8B** | single | 5 | 10 | 3,789.10 | 414.59 | 2,851.23 |
| **qwen_3_8B** | single | 10 | 10 | 3,070.20 | 412.86 | 2,136.25 |
| **qwen_3_8B** | single | 20 | 10 | 2,835.20 | 290.39 | 2,178.28 |
| **qwen_3_8B** | single | 40 | 10 | 2,722.60 | 310.32 | 2,020.60 |
| **qwen_3_8B** | single | 80 | 10 | 285,249.40 | 192,619.99 | -150,487.30 |

### Table S3. Total tokens by dataset × agent × batch size.

| dataset | type | N_tasks | n | mean_val | se_val | ci_low | ci_high |
|---|---|---|---|---|---|---|---|
| **gpt_mini** | multi | 5 | 10 | 3,744 | 226 | 3,231 | 4,256 |
| **gpt_mini** | multi | 10 | 10 | 7,935 | 244 | 7,383 | 8,486 |
| **gpt_mini** | multi | 20 | 10 | 14,875 | 217 | 14,384 | 15,366 |
| **gpt_mini** | multi | 40 | 10 | 30,330 | 382 | 29,465 | 31,195 |
| **gpt_mini** | multi | 80 | 10 | 60,371 | 275 | 59,750 | 60,992 |
| **gpt_mini** | single | 5 | 10 | 24,751 | 5,249 | 12,876 | 36,626 |
| **gpt_mini** | single | 10 | 10 | 159,246 | 32,061 | 86,720 | 231,772 |
| **gpt_mini** | single | 20 | 10 | 746,086 | 101,337 | 516,845 | 975,328 |
| **gpt_mini** | single | 40 | 10 | 1,595,768 | 360,754 | 779,686 | 2,411,850 |
| **gpt_mini** | single | 80 | 10 | 3,872,443 | 670,732 | 2,355,141 | 5,389,744 |
| **gpt_nano** | multi | 5 | 10 | 3,748 | 226 | 3,237 | 4,259 |
| **gpt_nano** | multi | 10 | 10 | 7,252 | 783 | 5,482 | 9,023 |
| **gpt_nano** | multi | 20 | 10 | 14,977 | 271 | 14,365 | 15,590 |
| **gpt_nano** | multi | 40 | 10 | 29,245 | 1,015 | 26,949 | 31,540 |
| **gpt_nano** | multi | 80 | 10 | 53,209 | 2,797 | 46,881 | 59,536 |
| **gpt_nano** | single | 5 | 10 | 14,652 | 1,415 | 11,450 | 17,854 |
| **gpt_nano** | single | 10 | 10 | 68,096 | 8,975 | 47,793 | 88,398 |
| **gpt_nano** | single | 20 | 10 | 113,612 | 29,759 | 46,291 | 180,932 |
| **gpt_nano** | single | 40 | 10 | 519,614 | 156,899 | 164,682 | 874,545 |
| **gpt_nano** | single | 80 | 10 | 891,986 | 512,762 | -267,962 | 2,051,934 |

| | | | | | | | |
|---|---|---|---|---|---|---|---|
| **llama_xlam_2_70b** | multi | 5 | 10 | 31,032 | 1,331 | 28,021 | 34,043 |
| **llama_xlam_2_70b** | multi | 10 | 10 | 70,094 | 2,265 | 64,970 | 75,217 |
| **llama_xlam_2_70b** | multi | 20 | 10 | 199,721 | 39,169 | 111,114 | 288,328 |
| **llama_xlam_2_70b** | multi | 40 | 10 | 370,747 | 54,371 | 247,751 | 493,744 |
| **llama_xlam_2_70b** | multi | 80 | 10 | 3,638,240 | 2,716,541 | -2,507,002 | 9,783,481 |
| **llama_xlam_2_70b** | single | 5 | 10 | 45,955 | 7,222 | 29,617 | 62,292 |
| **llama_xlam_2_70b** | single | 10 | 10 | 150,911 | 15,380 | 116,120 | 185,703 |
| **llama_xlam_2_70b** | single | 20 | 10 | 311,665 | 92,950 | 101,397 | 521,933 |
| **llama_xlam_2_70b** | single | 40 | 10 | 542,206 | 131,440 | 244,868 | 839,545 |
| **qwen_3_8B** | multi | 5 | 10 | 351,183 | 53,239 | 230,748 | 471,618 |
| **qwen_3_8B** | multi | 10 | 10 | 272,039 | 135,146 | -33,682 | 577,760 |
| **qwen_3_8B** | multi | 20 | 10 | 329,081 | 116,630 | 65,245 | 592,917 |
| **qwen_3_8B** | multi | 40 | 10 | 917,252 | 462,153 | -128,210 | 1,962,714 |
| **qwen_3_8B** | multi | 80 | 10 | 1,691,343 | 607,848 | 316,296 | 3,066,390 |
| **qwen_3_8B** | single | 5 | 10 | 3,898 | 477 | 2,819 | 4,976 |
| **qwen_3_8B** | single | 10 | 10 | 4,740 | 623 | 3,332 | 6,149 |
| **qwen_3_8B** | single | 20 | 10 | 4,516 | 250 | 3,949 | 5,083 |
| **qwen_3_8B** | single | 40 | 10 | 6,486 | 456 | 5,453 | 7,518 |
| **qwen_3_8B** | single | 80 | 10 | 161,092 | 104,749 | -75,866 | 398,050 |

Supplementary figures for the single model results.

**Figure 3. Token consumption by agent type and batch size**

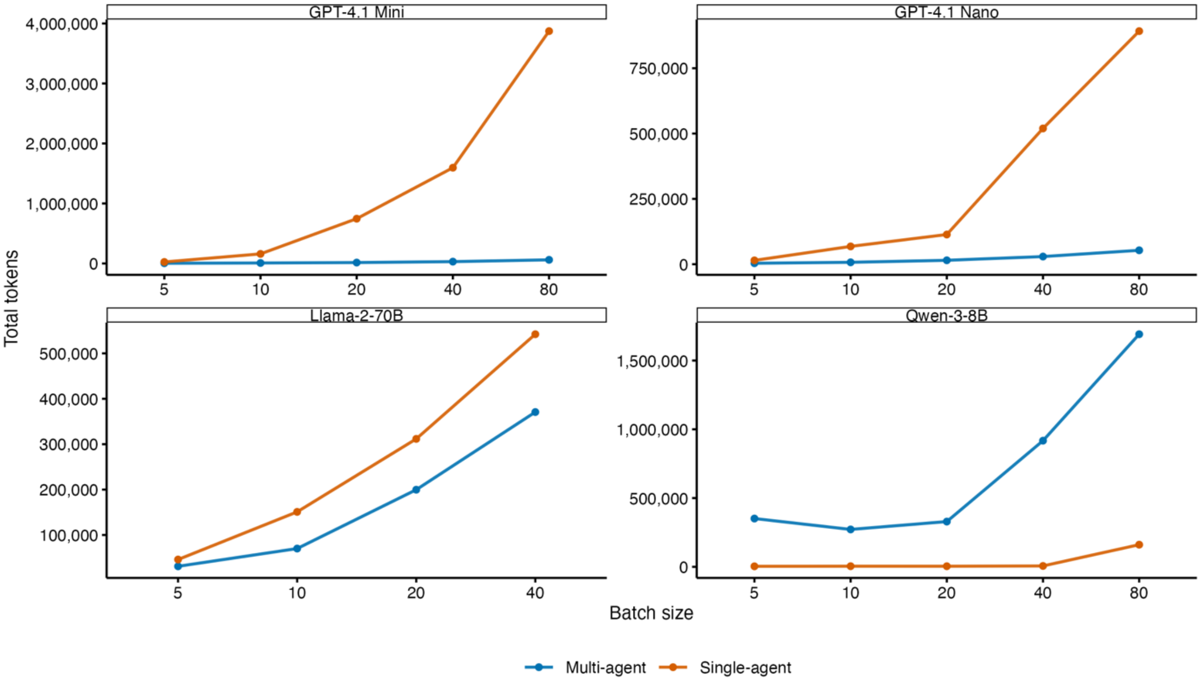


Token utilization patterns across different model architectures. Llama-2-70B data available up to batch size 40.

**Figure 2. Computational latency by agent type and batch size**

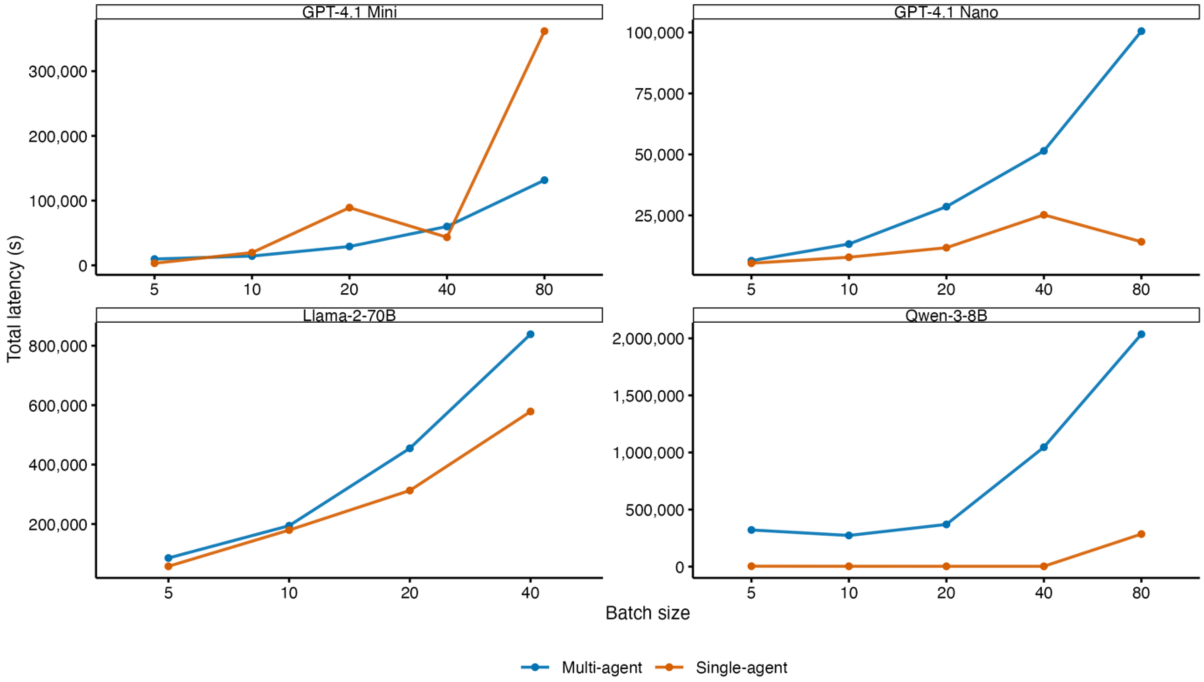


Latency scaling patterns vary across model architectures. Llama-2-70B evaluated up to batch size 40, other models up to 80.

**Figure 1. Task accuracy by agent type and batch size**

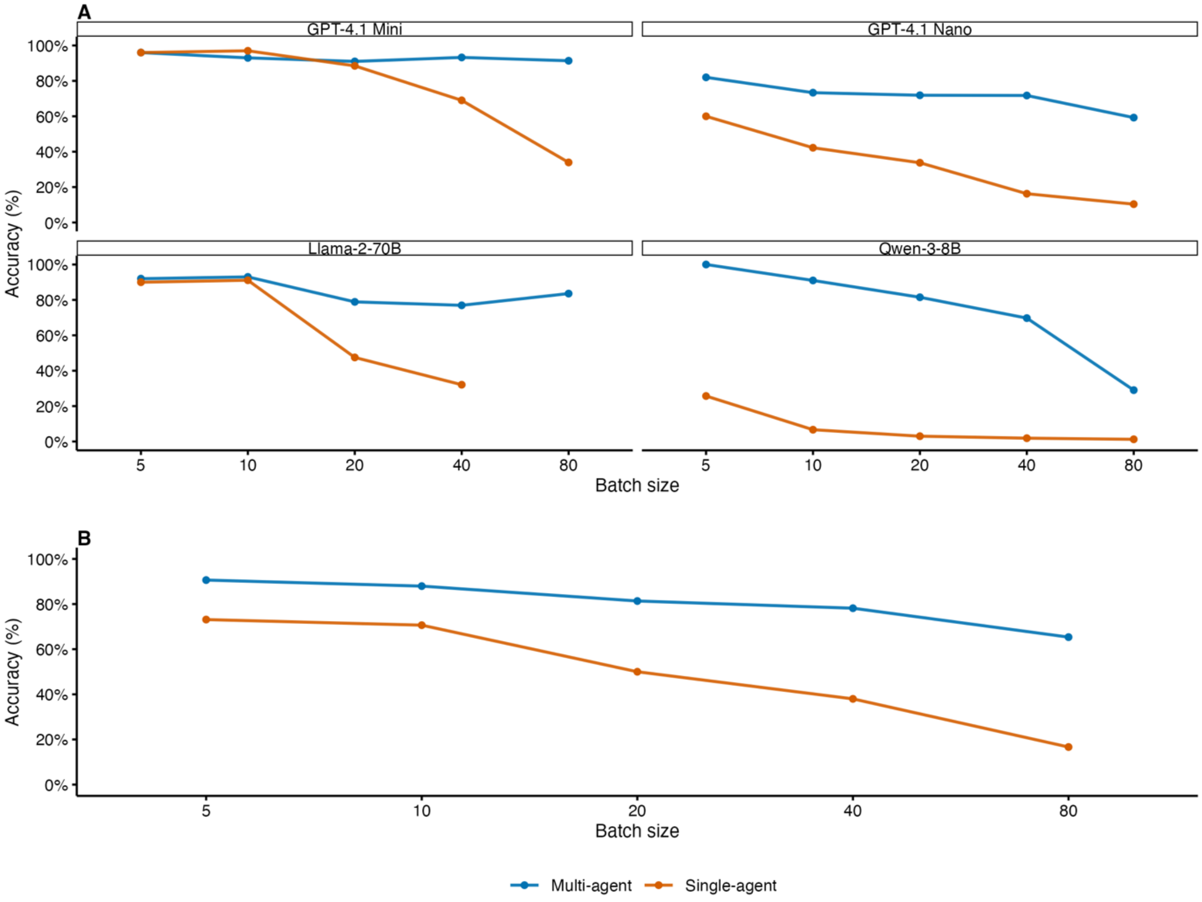


Multi-agent systems demonstrate superior accuracy retention across increasing batch sizes compared to single-agent approaches.

*Many single-agent runs, especially the larger batches for Llama-2-70B and Qwen-3-8B, terminated early with error JSON. These failures inflate single-agent latency and token counts in the comparison with the multi-agent runs.

**Section 2: Expanded methods.**

#### 1. *Data sources and ground truth*

We used three public corpora.

**Literature retrieval.** PubMed abstracts tagged *Neoplasms* and dated 2020-01-01 to 2025-05-30 (n = 234 650) were embedded with GIST-large (768 D, L2 normalised) and stored in a FAISS inner-product index.

**Clinical-note extraction.** All discharge summaries in MIMIC-IV that include the headers *Sex*, *Admission Date* and *Service* (n = 331 793) were retained. Admission dates were resampled uniformly across 2005-01-01 to 2019-12-31; discharge dates were offset 2–14 days later.

**Medication maths.** Twenty deterministic templates generated dosing questions that cover weight-based, body-surface-area, BMI, Cockcroft–Gault, infusion-rate and concentration conversions. Template parameters were drawn from uniform ranges.

Each task has one canonical answer string. Exact-match accuracy was computed after case-folding and whitespace normalisation.

#### 2. *Tools exposed to agents*

*Retriever* returns the *k* nearest abstracts plus cosine scores.

*Calculator* evaluates arithmetic expressions in a sandbox limited to Python's public *math* functions.

*Note fetcher* returns the full discharge summary for a given identifier.

#### 3. *Agent topologies*

*Single agent (SA).* One LLM receives the whole batch of *N* tasks and may use up to 10 × *N* dialogue turns.

*Multi agent (MA).* A lightweight orchestrator issues one *solve_task* call per task, passes the payload to a dedicated worker that may invoke one tool, and aggregates the worker outputs.

Four instruction-tuned checkpoints—GPT-4.1-mini, GPT-4.1-nano, Llama-2-70B-chat and Qwen-3-8B-chat—ran under both topologies. Random seed = 42.

#### 4. *Experimental design*

Batch sizes were 5, 10, 20, 40 and 80. Ten non-overlapping batches were generated for every (model × topology × batch size) cell. Open-weight models ran on one node with 8 × NVIDIA

H100-80 GB GPUs; hosted models used vendor APIs. Wall-clock latency was measured from the first model call to receipt of the final JSON. Total input and output tokens per batch were logged and divided by *N* for per-task values.

#### 5. *Statistical analysis*

Accuracy was compared with two-sided Welch t-tests. Total latency and token counts, which were skewed, used Wilcoxon rank-sum tests. P values were adjusted within each metric family by the Benjamini–Hochberg method. Analyses were performed in R 4.3.

**Section 3: Codes for running the agent pipelines.**
**Orchestrator pipeline**

```
--- Code Cell 1 ---
import json, ast, re, difflib, pandas as pd

def calculate_accuracy (result, df_tasks):

    try:

        agent_answers = ast.literal_eval (result)                # {"answer1": "...", ...}

        def clean(txt: object) -> str:
            """Lowercase, strip, drop quotes/commas/spaces."""
            return re.sub(r"[\"',]", "", str(txt)).strip().lower()

        def similar(a: str, b: str, thresh: float = 0.95) -> bool:
            """True if similarity ratio ≥ thresh."""
            ratio = difflib.SequenceMatcher(None, a, b).ratio()
            return ratio >= thresh

        results = []
        for i, gold in enumerate(df_tasks["gold_answer"], start=1):

            pred  = agent_answers.get(f"answer{i}", "")

            if pred.lower () == "male": pred = "M"

            if pred.lower () == "female": pred = "F"

            results.append (similar (clean(gold), clean(pred)))

        accuracy = sum(results) / len(results)

        return accuracy

    except:

        return "FAILED JSON"


--- Code Cell 2 ---
import pandas as pd
from pandas import NA
import time
```

```
def tag_and_accumulate(
    df: pd.DataFrame,
) -> pd.DataFrame:
    """
    Label every row and build cumulative-context columns, now INCLUDING
    each agent’s system prompt.

    Parameters
    ----------
    df : DataFrame with columns agent, role, content
    orch_system : str  (orchestrator's system prompt)
    worker_system : str  (worker system prompt, same for every worker)

    Returns
    -------
    DataFrame  (original + flag cols + orch_cumulative + worker_cumulative)
    """

    orch_system = """
You are an agent.

You will receive N independent clinical tasks.

For each task, call 'solve_task' (task) **one at a time**.

Only after all tasks are solved, return all the answers as:

{"answer1":"...", "answer2":"..." ... "answerN":"..."}

The answers must be just one term, no extra text, explanations or units."""

    worker_system = """
You are an agent.

You will recieve one task.

Choose one tool for this task:

• calculator(expression)
• pubmed_abstracts(query, top_k)
• note_text(note_id)

Return the answer as just one term, no extra text, explanations or units."""

    # ── basic masks
```

```
is_orch   = df["agent"] == "orchestrator"
is_worker = df["agent"] == "worker"
is_tool   = df["agent"] == "tool"

# ── worker-level flags ───────────────────────────────
worker_input  = (
    (is_worker & (df["role"] == "user")) |
    (is_tool   & df["role"].str.endswith(".output", na=False))
)

worker_output = (
    (is_worker & (df["role"] == "assistant")) |
    (is_tool   & df["role"].str.endswith(".input",  na=False))
)

# ── orchestrator flags ───────────────────────────────
orch_in  = (
    (is_orch & (df["role"] == "user")) |
    (is_worker & (df["role"] == "assistant"))
)

orch_out = (
    (is_orch & (df["role"] == "assistant")) |
    (is_worker & (df["role"] == "user"))
)

# ── worker index & propagation ───────────────────────
start_flags = is_worker & (df["role"] == "user")
df["worker"] = start_flags.cumsum()
df.loc[~(is_worker | is_tool), "worker"] = NA
df["worker"] = df["worker"].ffill()

# ── attach Boolean columns ───────────────────────────
df["orchestrator input"]  = orch_in.astype("boolean")
df["orchestrator output"] = orch_out.astype("boolean")
df["worker input"]        = worker_input.astype("boolean")
df["worker output"]       = worker_output.astype("boolean")

# ── cumulative context columns ───────────────────────
df["orch_cumulative"]   = NA
df["worker_cumulative"] = NA

# initialise the histories with the system prompts
orch_hist: list[str] = [orch_system] if orch_system else []
worker_hist: dict[int, list[str]] = {}
```

```
    for idx, row in df.iterrows():
        content = str(row["content"])

        # ─ orchestrator history grows on every row it sends/receives
        if is_orch[idx] or (is_worker[idx] and not is_tool[idx]):
            orch_hist.append(content)

        # ─ worker history (per-ID) grows on every worker/tool row
        wid = row["worker"]
        if pd.notna(wid):
            wid = int(wid)
            if wid not in worker_hist:
                # first time we meet this worker → start with system prompt
                worker_hist[wid] = [worker_system] if worker_system else []
            worker_hist[wid].append(content)

        # ─ snapshots taken only on *input* rows
        if orch_in[idx]:
            df.at[idx, "orch_cumulative"] = "\n".join(orch_hist)

        if worker_input[idx]:
            df.at[idx, "worker_cumulative"] = "\n".join(worker_hist[int(wid)])

    return df
```

```
--- Code Cell 3 ---
import tiktoken

def summarize_token_usage(df):
    """
    Print the total token count in each message category
    (orchestrator input/output, worker input/output).

    Assumes the DataFrame already has Boolean columns:
        • orchestrator input
        • orchestrator output
        • worker input
        • worker output
    and a `content` column holding the text.
    """
    enc = tiktoken.get_encoding("cl100k_base")  # GPT-4 family

    def token_sum(mask):
        return sum(len(enc.encode(str(text)))
```

```
                for text in df.loc[mask, "content"])

    totals = {
        "orch_in":   token_sum(df["orchestrator input"]),
        "orch_out":  token_sum(df["orchestrator output"]),
        "worker_in": token_sum(df["worker input"]),
        "worker_out":token_sum(df["worker output"]),
    }

    return totals
```

--- Code Cell 4 ---

```
import time
import asyncio
import nest_asyncio
from pathlib import Path
from datetime import datetime
import json
from typing import Any, Dict, List

import pandas as pd
import tiktoken
from openai import AsyncOpenAI
from agents import (
    Agent, Runner, OpenAIChatCompletionsModel, ModelSettings,
    function_tool, set_default_openai_client, set_default_openai_api,
    set_tracing_disabled
)

# ── logging helpers
```

---

```
encoder = tiktoken.get_encoding("cl100k_base")  # fallback encoding works for GPT-4 variants


def log_message(agent: str, role: str, content: str):
    """Record one message; write everything once, at the very end."""
    _LOG.append({
        "time":   datetime.utcnow().isoformat(timespec="seconds"),
        "agent":  agent,
        "role":   role,
        "tokens": len(encoder.encode(content)),
        "content": content,
```

```
    })

# ── data helpers
```

```
from calculator import Calculator
from pubmed_rag_index import SimplePubmedRAG


def build_openai_client():
    """Initialise the AsyncOpenAI client for Azure deployment."""
    nest_asyncio.apply()
    key_file = Path("azure openai research.txt")
    AZURE_KEY, AZURE_ENDPOINT = key_file.read_text().splitlines()[:2]

    # MODEL_DEPLOYMENT = "gpt-4o-mini-deploy-test-001"
    # MODEL_DEPLOYMENT = "gpt-41-nano-dzs-d3m-sandbox-swc-01-01"
    MODEL_DEPLOYMENT = "gpt-41-mini-dzs-d3m-sandbox-swc-01-01"
    # MODEL_DEPLOYMENT = "gpt-41-dzs-d3m-sandbox-swc-01-01"
    # MODEL_DEPLOYMENT = "gpt-o3-d3m-sandbox-clinicalrounding-swc-01-01"

    API_VERSION = "2024-12-01-preview"

    client = AsyncOpenAI(
        api_key=AZURE_KEY,

base_url=f"{AZURE_ENDPOINT.rstrip('/')}/openai/deployments/{MODEL_DEPLOYMENT}"
,
        default_headers={"api-key": AZURE_KEY},
        default_query={"api-version": API_VERSION},
    )
    set_default_openai_client(client, use_for_tracing=False)
    set_default_openai_api("chat_completions")
    set_tracing_disabled(True)
    return client, MODEL_DEPLOYMENT


client, MODEL_DEPLOYMENT = build_openai_client()

# ── load external data
```

```
df_notes = pd.read_pickle("notes/notes_10000.pkl")
df_abstracts = pd.read_pickle("pubmed/abstracts.pkl")
```

```
calc = Calculator()
rag = SimplePubmedRAG()
rag.load()

# ── domain tools with logging
────────────────────────────────────────────

@function_tool
def calculator(expression: str) -> str:
    """Return the numeric result of an arithmetic expression."""
    log_message ("tool", "calculator.input", expression)
    result = f"{calc.calculate(expression)['value']:.2f}"
    log_message ("tool", "calculator.output", result)
    return result


@function_tool
def pubmed_abstracts(query: str, top_k: int = 3) -> dict[str, str]:
    """Return {PMID: abstract_text} for the best matching PubMed hits."""
    log_message("tool", "pubmed_abstracts.input", query)
    hits = rag.search(query, top_k=top_k)
    abstracts = {
        str(h["PMID"]): df_abstracts.loc[
            df_abstracts["PMID"] == h["PMID"]
        ].iloc[0]["abstract"]
        for h in hits
    }
    log_message("tool", "pubmed_abstracts.output", json.dumps(abstracts)[:5000])  # safeguard
gigantic logs
    return abstracts


@function_tool
def note_text(note_id: str) -> str:
    """Fetch full clinical note text by note_id."""
    log_message("tool", "note_text.input", note_id)
    row = df_notes.loc[df_notes["note_id"] == note_id]
    text = "" if row.empty else row.iloc[0]["text"]
    log_message("tool", "note_text.output", text[:5000])
    return text

# ── worker agent
────────────────────────────────────────────────────────────

worker_agent = Agent(
    name="Clinical-Worker",
```

```
    instructions=(
"""
You are an agent.

You will recieve one task.

Choose one tool for this task:

• calculator(expression)
• pubmed_abstracts(query, top_k)
• note_text(note_id)

Return the answer as just one term, no extra text, explanations or units.
"""
    ),
    model=OpenAIChatCompletionsModel(model=MODEL_DEPLOYMENT,
openai_client=client),
    model_settings=ModelSettings(),
    tools=[calculator, pubmed_abstracts, note_text],
)

# ── expose the worker as a tool, with logging ──────────────────────

def make_worker_tool(agent: Agent):
    @function_tool(
        name_override="solve_task",
        description_override="Solve a single clinical task and return the answer."
    )
    async def solve_task(task: str) -> str:

        log_message("worker", "user", task)

        result = await Runner.run (agent, task, max_turns=10)

        # ── worker's answer (output) ──
        log_message("worker", "assistant", result.final_output)

        return result.final_output

    return solve_task

solve_task = make_worker_tool(worker_agent)

# ── orchestrator agent
```

```
orchestrator_agent = Agent(
    name="Orchestrator",
    instructions=(
"""
You are an agent.

You will receive N independent clinical tasks.

For each task, call 'solve_task' (task) **one at a time**.

Only after all tasks are solved, return all the answers as:

{"answer1":"...", "answer2":"..." ... "answerN":"..."}

The answers must be just one term, no extra text, explanations or units.
"""
    ),
    model=OpenAIChatCompletionsModel(model=MODEL_DEPLOYMENT,
openai_client=client),
    model_settings=ModelSettings(parallel_tool_calls=False),
    tools=[solve_task],
)

# ── helper to run a batch of tasks
─────────────────────────────────────

def run_batch(tasks: List[str], max_turns: int = 200):

    composite_prompt = (
        "Here are the tasks. Solve them one by one:\n\n" +
        "\n".join(f"{i+1}. {t}" for i, t in enumerate(tasks))
    )

    log_message("orchestrator", "user", composite_prompt)

    result = Runner.run_sync (orchestrator_agent, composite_prompt, max_turns=max_turns)

    # log the orchestrator's reply
    log_message("orchestrator", "assistant", result.final_output)

    return result

# ── example entry point
───────────────────────────────────────────────

df_all = pd.read_pickle("tasks_set.pkl")
```

```
_LOG = []

offset = 0                    # running position in df_all

for N_tasks in [5, 10, 20, 40, 80]:

    for exp in range(1, 11):      # 1-based for nicer file names

        _LOG.clear()            # start fresh

        df_tasks = df_all.iloc [offset:offset + N_tasks]

        offset += N_tasks        # advance for next run

        stem = f"multi_{MODEL_DEPLOYMENT}_{N_tasks}_{exp}"

        # Ensure the directory exists; create it if it does not
        Path(f"singles/{MODEL_DEPLOYMENT}").mkdir(parents=True, exist_ok=True)

        log_path = Path(f"singles/{MODEL_DEPLOYMENT}") / f"{stem}_log.pkl"

        if log_path.exists():

            print (f"{log_path} already exists – skipping round")

            continue

        start = time.perf_counter()
        result = run_batch (df_tasks["prompt"].tolist())
        latency_ms = int ((time.perf_counter() - start) * 1000)

        df = pd.DataFrame(_LOG)

        df = tag_and_accumulate (df)

        df.to_pickle(f"singles/{MODEL_DEPLOYMENT}/{stem}_log.pkl")

        df_tasks.to_pickle(f"singles/{MODEL_DEPLOYMENT}/{stem}_tasks.pkl")

        accuracy = calculate_accuracy (result.final_output, df_tasks)

        totals = summarize_token_usage (df)

        df_summarize = pd.DataFrame ({
            "offset":[offset],
```

```
    "latency":[latency_ms],
    "model":[MODEL_DEPLOYMENT],
    "exp":[exp],
    "N_tasks":[N_tasks],
    "accuracy":[accuracy],
    "input_tokens" : [totals ["orch_in"] + totals ["worker_in"]],
    "output_tokens" : [totals ["orch_out"] + totals ["worker_out"]],
    "orchestrator_input_tokens" : [totals ["orch_in"]],
    "orchestrator_output_tokens" : [totals ["orch_out"]],
    "worker_input_tokens" : [totals ["worker_in"]],
    "worker_output_tokens" : [totals ["worker_out"]]
})

df_summarize.to_pickle (f"singles/{MODEL_DEPLOYMENT}/{stem}_summarize.pkl")

print (stem, latency_ms, accuracy, totals)
```

**Singel agent pipeline:**

```
--- Code Cell 1 ---
import json, ast, re, difflib, pandas as pd

def calculate_accuracy (result, df_tasks):

    try:

        agent_answers = ast.literal_eval (result)                # {"answer1": "...", ...}

        def clean(txt: object) -> str:
            """Lowercase, strip, drop quotes/commas/spaces."""
            return re.sub(r"[\"',]", "", str(txt)).strip().lower()

        def similar(a: str, b: str, thresh: float = 0.95) -> bool:
            """True if similarity ratio ≥ thresh."""
            ratio = difflib.SequenceMatcher(None, a, b).ratio()
            return ratio >= thresh

        results = []

        for i, gold in enumerate (df_tasks["gold_answer"], start=1):

            pred  = agent_answers.get(f"answer{i}", "")

            if pred.lower () == "male": pred = "M"

            if pred.lower () == "female": pred = "F"

            results.append (similar (clean(gold), clean(pred)))

        accuracy = sum(results) / len(results)

        return accuracy

    except:

        return "FAILED JSON"


--- Code Cell 2 ---

import pandas as pd
import tiktoken
```

```
def enrich_chat_log (df: pd.DataFrame,
                 system_prompt: str = "this is system prompt") -> pd.DataFrame:
    """
    Adds chat-aware token-count columns to a conversation trace.

    New columns
    -----------
    input_tokens  – tokens in the current row that the model receives
    output_tokens – tokens in the current row that the model emits
    chat          – total tokens in the running chat after this row
    real_input    – chat for input rows, 0 for output rows
    """
    enc = tiktoken.get_encoding("cl100k_base")
    sys_tokens = len(enc.encode(system_prompt))

    def n_tokens(txt) -> int:
        return len(enc.encode(str(txt)))

    def classify(row):
        """Return (in_tok, out_tok, is_input_row)."""
        if row.get("role") in ("user", "tool") or row.get("type") == "function_call_output":
            txt = row.get("content") if pd.notnull(row.get("content")) else row.get("output")
            t = n_tokens(txt)
            return t, 0, True
        txt = row.get("content") if pd.notnull(row.get("content")) else row.get("arguments")
        t = n_tokens(txt)
        return 0, t, False


    cols = df.apply(classify, axis=1, result_type="expand")

    cols.columns = ["input_tokens", "output_tokens", "is_input"]

    df = pd.concat([df, cols], axis=1)

    chat_tokens = sys_tokens
    chat_list, real_input_list = [], []

    for in_tok, out_tok, is_in in df[["input_tokens", "output_tokens",
"is_input"]].itertuples(index=False):
        if is_in:
            chat_tokens += in_tok
            real_input_list.append(chat_tokens)
        else:
            chat_tokens += out_tok
            real_input_list.append(0)
```

```
        chat_list.append(chat_tokens)

    df["chat"] = chat_list

    df["real_input"] = real_input_list

    return df


--- Code Cell 3 ---
"""
Single-agent stress test: clinical agent → three tools
Revised: give the agent many tasks at once; it answers them sequentially
and returns a JSON array of answers.
"""

import time
import nest_asyncio
import pandas as pd
from pathlib import Path
from openai import AsyncOpenAI
from agents import (                    # same SDK import block
    Agent, Runner, OpenAIChatCompletionsModel, ModelSettings,
    function_tool, set_default_openai_client, set_default_openai_api,
    set_tracing_disabled,
    ItemHelpers, MessageOutputItem, ToolCallItem,
    ToolCallOutputItem, ReasoningItem,
)

# ── initial setup identical to your original file ────────────────────────
nest_asyncio.apply()
key_file = Path("azure openai research.txt")
AZURE_KEY, AZURE_ENDPOINT = key_file.read_text().splitlines()[:2]

# MODEL_DEPLOYMENT = "gpt-4o-mini-deploy-test-001"
# MODEL_DEPLOYMENT = "gpt-41-nano-dzs-d3m-sandbox-swc-01-01"
MODEL_DEPLOYMENT = "gpt-41-mini-dzs-d3m-sandbox-swc-01-01"
# MODEL_DEPLOYMENT = "gpt-41-dzs-d3m-sandbox-swc-01-01"

API_VERSION      = "2024-12-01-preview"

client = AsyncOpenAI(
    api_key=AZURE_KEY,

base_url=f"{AZURE_ENDPOINT.rstrip('/')}/openai/deployments/{MODEL_DEPLOYMENT}"
,
```

```
    default_headers={"api-key": AZURE_KEY},
    default_query={"api-version": API_VERSION},
)
set_default_openai_client(client, use_for_tracing=False)
set_default_openai_api("chat_completions")
set_tracing_disabled(True)

# ── external data and tools (unchanged) ─────────────────────────────────

df_notes     = pd.read_pickle("notes/notes_10000.pkl")
df_abstracts = pd.read_pickle("pubmed/abstracts.pkl")

from calculator import Calculator
from pubmed_rag_index import SimplePubmedRAG
rag = SimplePubmedRAG(); rag.load()
calc = Calculator()

@function_tool
def calculator(expression: str) -> str:
    """Return the numeric result of an arithmetic expression."""
    return str(calc.calculate(expression)["value"])

@function_tool
def pubmed_abstracts(query: str, top_k: int = 3) -> dict[str, str]:
    """Return {PMID: abstract_text} for the best matching PubMed hits."""
    hits = rag.search(query, top_k=top_k)
    return {
        str(h["PMID"]): df_abstracts.loc[
            df_abstracts["PMID"] == h["PMID"]
        ].iloc[0]["abstract"]
        for h in hits
    }

@function_tool
def note_text(note_id: str) -> str:
    """Fetch full clinical note text by note_id."""
    row = df_notes.loc[df_notes["note_id"] == note_id]
    return "" if row.empty else row.iloc[0]["text"]

# ── clinical agent ───────────────────────────────────────────────────────

clinical_agent = Agent(
    name="Clinical-Agent",
    instructions=(
"""
You are an agent.
```

```
You will receive N independent tasks at once.

Solve them sequentially, one by one using the tools:

• calculator(expression)
• pubmed_abstracts(query, top_k)
• note_text(note_id)

When you have all the answers, return in the format:

{"answer1": <"your answer">, "answer2":"<your answer>"...]}

Each answer must be just one term, without extra text, explanations, units, etc.
"""
   ),
   model=OpenAIChatCompletionsModel(model=MODEL_DEPLOYMENT,
openai_client=client),
   model_settings=ModelSettings(),
   tools=[calculator, pubmed_abstracts, note_text],
)


# ── load tasks and build one composite prompt


df_all = pd.read_pickle("tasks_set.pkl")

offset = 0                    # running position in df_all

for N_tasks in [5, 10, 20, 40, 80]:

   for exp in range (1, 11): # range(1, 6):      # 1-based for nicer file names

      df_tasks = df_all.iloc [offset:offset + N_tasks]

      offset += N_tasks        # advance for next run

      stem = f"single_{MODEL_DEPLOYMENT}_{N_tasks}_{exp}"

      # Ensure the directory exists; create it if it does not
      Path(f"singles/{MODEL_DEPLOYMENT}").mkdir(parents=True, exist_ok=True)

      log_path = Path(f"singles/{MODEL_DEPLOYMENT}") / f"{stem}_log.pkl"
```

```
if log_path.exists():

    print (f"{log_path} already exists – skipping round")

    continue

multi_task_prompt = (
    "Here are the tasks. Solve them one by one:\n\n" +
    "\n".join(f"{i+1}. {t}" for i, t in enumerate(df_tasks["prompt"].tolist()))
)

t0       = time.monotonic()

result   = Runner.run_sync (clinical_agent, multi_task_prompt,
                             max_turns = 10 * N_tasks)

msgs = result.to_input_list()          # list of plain dicts

df_log = pd.json_normalize(msgs)

df_log = enrich_chat_log (df_log)

latency  = int((time.monotonic() - t0) * 1000)


df_log.to_pickle(f"singles/{MODEL_DEPLOYMENT}/{stem}_log.pkl")

df_tasks.to_pickle(f"singles/{MODEL_DEPLOYMENT}/{stem}_tasks.pkl")

accuracy = calculate_accuracy (result.final_output, df_tasks)

df_summarize = pd.DataFrame ({
    "offset":[offset],
    "latency":[latency],
    "model":[MODEL_DEPLOYMENT],
    "exp":[exp],
    "N_tasks":[N_tasks],
    "accuracy":[accuracy],
    "input_tokens" : [df_log ["real_input"].sum ()],
    "output_tokens" : [df_log ["output_tokens"].sum ()],
})


df_summarize.to_pickle (f"singles/{MODEL_DEPLOYMENT}/{stem}_summarize.pkl")

print (stem, latency, accuracy,
```

```
df_log ["real_input"].sum (),
df_log ["output_tokens"].sum ())
```
